# Diabetogenic elevated childhood total fat in South Asian and Black African/Caribbean people relates to adverse early life growth and low socioeconomic position compared to White people in the UK

**DOI:** 10.1101/2025.02.12.25322137

**Authors:** Kishan Patel, Sophie V Eastwood, Jonathan C Wells, Nish Chaturvedi, Charis Bridger-Staatz

## Abstract

**Aims/hypothesis:** Excess type 2 diabetes mellitus in ethnic minorities remains unexplained, though greater fat mass makes a strong contribution. We hypothesized that height and weight through infancy in South Asian and Black African/Caribbean subgroups is more adverse than in White populations. These, allied to poor socioeconomic position, determine greater fat mass at age 7.

**Methods:** We report a secondary analysis from the UK Millennium Cohort Study, including 12280 White, 358 Indian, 650 Pakistani, 268 Bangladeshi, 163 Black Caribbean and 277 Black African births between 2000-2002. Birthweight was reported, and heights and weights measured at ages 3, 5, 7, 11, 14, and 17. Bioimpedence captured fat mass, indexed to weight, at ages 7, 11, 14 and 17. Standardised differences in anthropometry, using Whites as the comparator, were calculated. We explored the effect of early growth on ethnic differences in fat mass index at age 7. Confounders included maternal anthropometry, smoking, infant breastfeeding, education, parental income and area level socioeconomic deprivation.

**Results:** All ethnic minority subgroups had lower birthweight and accelerated infant height and weight growth compared to White children. By age 3, mean height was greater in all ethnic minorities than in White children. This height advantage was progressively lost, first in Bangladeshi children. By age 17 in boys and girls, Indians were 1.8/2.5 cm, Pakistanis 2.2/3.4 cm, Bangladeshis 4.8/6.0 cm, and Black Caribbeans 1.6/0.5 cm shorter than White children. Heights were equivalent in Black African children. By age 17, all South Asians were lighter, and Black African/Caribbeans heavier than White children. The anthropometric gradient by ethnicity in children mirrored that in mothers. Ethnic minority girls were more likely to be menstruating by age 11 than White girls (range 12-27% versus 9%). At age 7, standardized fat mass index (kg/m^2^) in boys/girls was 0.17/0.01 standard deviations greater in Indian, 0.21/0.04 in Pakistani, 0.18/0.16 in Bangladeshi, 0.48/0.35 in Black Caribbean, and 0.37/0.75 in Black African children compared to White children. These persisted to age 17. Weight gain to age 3, and in Black Africans/Caribbeans, adverse individual and neighbourhood socioeconomic position contributed to accounting for ethnic differences in fat mass.

**Conclusions/Interpretation:** Ethnic minorities in the UK have poorer childhood growth than White children, achieving shorter height, greater fat mass and early female puberty. Mirroring of maternal and offspring ethnic subgroup gradients in height and weight indicates inter-generational transmission. Persistent adverse socioeconomic circumstances perpetuate ethnic adversity in early life accrual of body fat.

**Research in context:** *What is already known about this subject?:*

- Ethnic minority groups have early and excess risks of type 2 diabetes compared to Whites
- Ethnic minorities are known to have lower birthweight, and experience accelerated infant growth.
- Adult fat mass is greater in ethnic minority groups

*What is the key question?:*

- Can ethnic differences in early growth, maternal body size, child rearing practices and socioeconomic position account for ethnic differences in child fat mass and fat free mass?

*What are the new findings?:*

- All ethnic minority subgroups experience low birthweight and accelerated infant growth, and all, bar Black African girls, are shorter by age 17 compared to Whites.
- The magnitude of difference in achieved height and weight varies markedly by ethnic subgroup and mirrors the ethnic gradient observed in mothers.
- Accelerated infant growth contributes to excess childhood fat mass in children of Indian, Pakistani, Bangladeshi, Black African and Black Caribbean descent. Adverse individual and neighbourhood socioeconomic status makes an additional contribution in Black African and Black Caribbean children.

*How might this impact on clinical practice in the future?:*

- Resolving parental and childhood individual and area socioeconomic inequalities is critical to reducing adverse early growth and excess adiposity that predisposes to type 2 diabetes.

## Introduction

People of South Asian and of African (Black African and Black Caribbean) ethnicity have markedly greater risks and earlier age of onset of type 2 diabetes mellitus than White Europeans^1,2^. Explanations for this premature excess are not entirely understood, although greater total body fat, and adverse fat distribution are major contributors^2^. Poor in utero development, proxied by low birthweight, coupled with adverse patterns of accelerated early post-natal growth are associated both with elevated total body fat and with type 2 diabetes mellitus associated traits at birth and in childhood, and likely make a strong contribution to premature disease onset^3,4^. Low birthweight and accelerated early height growth have been reported overall in both South Asians and African Caribbeans^5^, but not by ethnic subgroup and not in association with fat mass. Separately, while low fat free mass is also evident in South Asians^6^, making an additional contribution to type 2 diabetes mellitus risk^7^, lean mass appears greater in people of African descent^6^. These previous studies did not have historical growth data so associations between growth and fat mass could not be explored. Additionally, central fat deposition, viewed as a hallmark of adverse adiposity patterns in South Asians and also contributing to excess type 2 diabetes mellitus, is not always consistently observed^1^ and does not appear to feature in people of African descent^2^.

Whilst biology has been invoked to account for ancestral differences in body habitus and type 2 diabetes mellitus risk, socioeconomic factors also strongly influence differential growth and fat distribution patterns. Socioeconomic status (SES) has had a marked impact on secular trends in height and weight^8^, and both individual and neighbourhood measures of socioeconomic position determine divergent fat mass trajectories^9^. The more adverse SES of ethnic minorities in the UK strongly contributes to lower birthweights compared to the White population^10^. Both ethnicity and SES have also been associated with earlier age at menarche, itself associated with adiposity^11^.

We hypothesized that early life growth patterns in the UK differ by ethnic subgroup. People of South Asian and African ethnicity will be shorter and heavier by late adolescence compared to Whites, and pubertal maturation will occur earlier. While both ancestries will have greater fat mass than Whites, fat free mass will be lower in South Asians, but higher in children of Black Caribbean and Black African descent. Ethnic subgroups, by parental country of origin, will have different growth patterns in association with differences in SES. These different growth patterns, coupled with adverse socioeconomic circumstances, will account for ethnic differences in both fat mass and fat free mass in childhood.

## Methods

These are secondary analyses of the Millennium Cohort Study (MCS). This is a UK nationally representative birth cohort, oversampling regions with higher concentrations of ethnic minority populations. It consists of 19,244 families who had children born in the UK between the years 2000 and 2002. Recruitment was at 9 months of age of the child for 18,552 families, and 3 years of age for 692 families. Questionnaires to parents captured information around birth, and were repeated when children were ages 3, 5, 7, and 11. At age 14, both the child and parents provided information, and at age 17, the young people responded about themselves. Ethics approval was obtained by the National Health Service Research Ethics Committee up to age 14 years, and the National Research Ethics Service at age 17 years. Informed parental consent was obtained in advance of data collection up to age 14 years. At age 17 years, informed verbal consent was obtained by the participant. Participants were able to refuse completing any element of the study or withdraw at any time, even at younger ages when written consent was only obtained from the parent.

Parents were asked to assign ethnicity of their children using the UK census groupings of: White, Indian, Pakistani, Bangladeshi, Black Caribbean, and Black African^12^. Specific mixed ethnicity groups (eg Black Caribbean/White, Pakistani/White) or other ethnicities were small and therefore excluded from this analysis.

The main outcome variables were height from age 3 to age 17, weight including birthweight to age 17, and Fat Mass Index (FMI), and Fat-Free Mass Index (FFMI) at age 7, 11, 14 and 17. Anthropometric measures have been described^9^; briefly, body fat percentage (FM%) was collected by foot-to-foot bio-electrical impedance analysis (BIA) using the Tanita (Bf-522W; Tokyo, Japan) scales, performed by trained interviewers following standardised protocols. Equations used to calculate FM% were those used by the manufacturer. Height and weight were measured before BIA assessment, and inputted into the scales to enable output of FM%. Fat mass (FM) was calculated by dividing FM% by 100 and multiplying by total body weight ((FM = (FM%/100) × weight)). FMI was derived by indexing FM to height squared (FMI = FM/height²). FFM was calculated by subtracting FM from total body weight (FFM = weight - FM), and FFMI was obtained by indexing FFM to height squared (FFMI = FFM/height²). The FMI/FFMI ratio at age 7 was calculated. In girls, onset of menstruation by the time of the age 11 questionnaire was used as a proxy for puberty, as previously reported^13^.

Mediators accounting for differences between ethnicities in FMI and FFMI at age 7 included the growth measures of birthweight, relative weight gain from birth to age 3, (calculated by subtracting birth weight from weight at age 3, and dividing by birthweight in kg, this captures the early post- natal accelerated growth period), waist circumference at ages 5 and 7, and waist/height ratio at ages 5 and 7. Maternal factors included self-reported maternal height and pre-pregnancy weight, and calculated body mass index (BMI), smoking status and breastfeeding duration to 4 months captured at recruitment. Socioeconomic position was assessed using the Index of Multiple Deprivation (IMD), an area measure of deprivation at birth, as well as individual measures including parental education (college degree/no college degree), and highest parental income quintiles at birth of the study participant. Ethics approval for data collection was granted by the NHS Research Ethics Committee up to age 14, and then by the National Research Ethics Service at age 17 years.

### Statistical Analyses

The analytic sample comprised all cohort members with complete information on ethnicity and at least one response for each outcome and mediator from birth to age 17. Sample design weights were used to correct for MCS cases having unequal probabilities of selection that result from the stratified cluster sample design. The data were survey-weighted, with MCS sampling weights applied to each statistical model to ensure the results accurately represent the UK population^14^.

Multiple imputation was performed to address missing data on mediators, under a missing at random assumption. Twenty imputations were generated, accounting for item nonresponse and dropout. Birthweight-related variables and ethnicity were included in the imputation model as predictors of missingness^15^. This approach ensures that variability and uncertainty associated with the missing data are properly accounted for, leading to more robust and reliable statistical analyses, preserving variable relationships, and minimising bias.

Descriptive analyses calculated averages and percentages of maternal characteristics, parental sociodemographic indices, and early life anthropometric measures, stratified by ethnicity and sex. Standardised measures of height, weight, FM and FFM were calculated at each age using the standard deviation of the White group.

Regression analyses calculated coefficients for FMI and FFMI, comparing ancestral groups to Whites at age 7, (the first time these were measured), stratified by sex. Three models were employed: Model 1 - unadjusted, Model 2 - adjusted for early life growth, ie weight gain from birth to age three, and Model 3 - fully adjusted for maternal, early life, and SES factors.

All analyses were performed using STATA 18. Graphs were generated using R.

## Results

### Demographic characteristics of the sample by ethnicity

Of the 18,786 mothers of participants who had any interaction with the ethnicity module at recruitment, 48 did not complete it. A further 562 were excluded as being of “mixed ethnicity”, and 308 as being “other ethnicity”. The remaining 17,868 recruits ethnicity was as follows; White - 15,491, Indian – 470, Pakistani – 903, Bangladeshi – 368, Black Caribbean – 247 and Black African – 389. From there, exclusions were only on the basis of not having at least one data point for our outcomes. The ethnicity of those remaining was: Whites – 12280, Indian – 358, Pakistani – 650, Bangladeshi - 268, Black Caribbean - 163, and Black African 277 (table 1).

**Table 1:** Maternal characteristics, parental sociodemographic indices and offspring birthweight, early life weight gain, waist to height ratio, FMI/FFMI ratio and in girls, early puberty, by ethnicity (mean ± SD, or %)

|  | White<br>*n*=12280 | Indian<br>*n*=358 | Pakistani<br>*n*=650 | Bangladeshi<br>*n*=268 | Black Caribbean<br>*n*=163 | Black African<br>*n*=277 |
| --- | --- | --- | --- | --- | --- | --- |
| Maternal height (m) | 1.64 ± 0.08 | 1.60 ± 0.08 | 1.61 ± 0.08 | 1.57 ± 0.09 | 1.65 ± 0.08 | 1.65 ± 0.10 |
| Maternal weight (kg) | 64.2 ± 15.0 | 56.8 ± 13.4 | 60.8 ± 14.7 | 58.5 ± 15.6 | 67.9 ± 16.9 | 68.3 ± 18.5 |
| Maternal BMI (kg/m<sup>2</sup>) | 23.7 ± 5.3 | 22.3 ± 5.1 | 23.5 ± 5.9 | 23.7 ± 6.4 | 25.0 ± 5.7 | 25.1 ± 6.3 |
| Maternal smoking % | 12.5 | 4.1 | 3.6 | 2.3 | 9.6 | 1.8 |
| Breast fed to 4 months % | 22.6 | 25.6 | 14.5 | 19.2 | 39.5 | 39.8 |
| Area deprivation (IMD*) | 5.8 ± 3.2 | 5.0 ± 3.9 | 2.7 ± 3.7 | 2.5 ± 3.7 | 3.4 ± 2.8 | 3.0 ± 3.2 |
| Degree level education % | 29.7 | 37.8 | 14.2 | 12.3 | 30.4 | 40.3 |
| Highest parental income quintile at birth % | 18.8 | 17.8 | 3.7 | 2.2 | 9.8 | 9.0 |
| Boys - *n* | **6274** | **184** | **330** | **129** | **85** | **147** |
| Birthweight (kg) | 3.46 ± 0.69 | 3.03 ± 0.92 | 3.15 ± 0.66 | 3.13 ± 0.59 | 3.27 ± 0.90 | 3.37 ± 0.77 |
| Relative weight gain birth to age 3 | 4.57 ± 1.37 | 5.14 ± 2.56 | 4.87 ± 1.30 | 4.93 ± 1.38 | 5.20 ± 2.40 | 4.98 ± 1.09 |
| Waist circumference (cm) age 5 | 54.01 ±<br>4.50 | 53.17 ± 7.03 | 52.58 ±<br>6.03 | 53.52 ± 6.22 | 55.26 ± 6.66 | 53.98 ± 5.57 |
| Waist/height ratio age 5 | 0.49 ± 0.04 | 0.47 ± 0.06 | 0.47 ± 0.05 | 0.48 ± 0.05 | 0.49 ± 0.06 | 0.47 ± 0.04 |
| Waist circumference (cm) age 7 | 57.13 ±<br>6.18 | 57.31 ± 8.27 | 57.10 ±<br>8.89 | 57.04 ± 7.80 | 58.75 ± 9.16 | 58.01 ± 7.40 |
| Waist/height ratio age 7 | 0.46 ± 0.04 | 0.46 ± 0.06 | 0.46 ± 0.06 | 0.46 ± 0.05 | 0.47 ± 0.07 | 0.45 ± 0.04 |
| FMI/FFMI ratio age 7 | 0.25 ± 0.09 | 0.28 ± 0.12 | 0.28 ± 0.14 | 0.27 ± 0.10 | 0.29 ± 0.15 | 0.29 ± 0.10 |
| Girls – *n* | **6006** | **171** | **320** | **139** | **78** | **130** |
| Birthweight (kg) | 3.34 ± 0.66 | 2.96 ± 0.81 | 3.08 ± 0.67 | 2.97 ± 0.95 | 3.15 ± 0.72 | 3.16 ± 0.72 |
| Relative weight gain birth to age 3 | 4.52 ± 1.37 | 5.01 ± 2.22 | 4.94 ± 1.71 | 5.29 ± 3.56 | 5.41 ± 3.64 | 5.36 ± 1.63 |
| Waist circumference (cm) age 5 | 53.78 ±<br>4.80 | 52.75 ± 5.67 | 53.11 ±<br>5.82 | 53.68 ± 7.62 | 54.88 ± 5.13 | 55.72 ± 6.35 |
| Waist/height ratio age 5 | 0.49 ± 0.04 | 0.47 ± 0.05 | 0.48 ± 0.05 | 0.49 ± 0.07 | 0.49 ± 0.04 | 0.49 ± 0.05 |
| Waist circumference (cm) age 7 | 57.03 ±<br>6.55 | 56.51 ± 7.30 | 57.07 ±<br>7.92 | 56.95 ± 8.92 | 58.49 ± 7.07 | 61.31 ± 9.70 |
| Waist height ratio age 7 | 0.46 ± 0.05 | 0.46 ± 0.05 | 0.46 ± 0.06 | 0.46 ± 0.06 | 0.46 ± 0.05 | 0.48 ± 0.06 |
| FMI/FFMI ratio age 7 | 0.29 ± 0.11 | 0.30 ± 0.11 | 0.30 ± 0.12 | 0.31 ± 0.14 | 0.32 ± 0.12 | 0.37 ± 0.16 |
| Menstruation onset before age 11 % | 9.0 | 26.9 | 11.8 | 27.2 | 17.9 | 22.8 |

\*IMD = index of multiple deprivation

ESM table 1 shows missing data, ranging from 1 for index of multiple deprivation, to 6118 for fat mass at age 17 years. South Asian mothers were shorter and lighter than their White counterparts, whereas Black Caribbean and Black African mothers were of similar height but heavier (Table 1). Breastfeeding for at least four months was highest among Black Caribbean and Black African mothers. All ethnic minority group mothers were less likely to smoke than White European mothers. Indian, Black Caribbean and Black African mothers were equally or more likely to be educated to degree level compared to White mothers. In contrast, all minority groups were less likely to be in the highest quintile of parental income, and more likely to live in a socioeconomically deprived neighbourhood than their White counterparts. For the latter, Bangladeshi families were most disadvantaged, followed by Pakistani, Black African, Black Caribbean and Indian families.

### Weight at birth through to age 17 by ethnicity

Boys and girls from all minority groups had lower birthweights compared to White boys and girls, with the most marked differences observed in Indian children (standardized difference: -0.62 for boys, -0.58 for girls), and least in Black African children (-0.13 for boys, -0.27 for girls) (Table 1, Figure 1, ESM tables 2a and 2b). Relative weight gain to age 3 was greater in all minority groups compared to Whites (table 1). Black Caribbean and Black African children were heavier at age 3, and remained so to age 17, compared to White children. In contrast, South Asian children were lighter than White children at all ages, except briefly for boys at age 11 (Figure 1, ESM Tables 2a and 2b).

**Figure 1.**
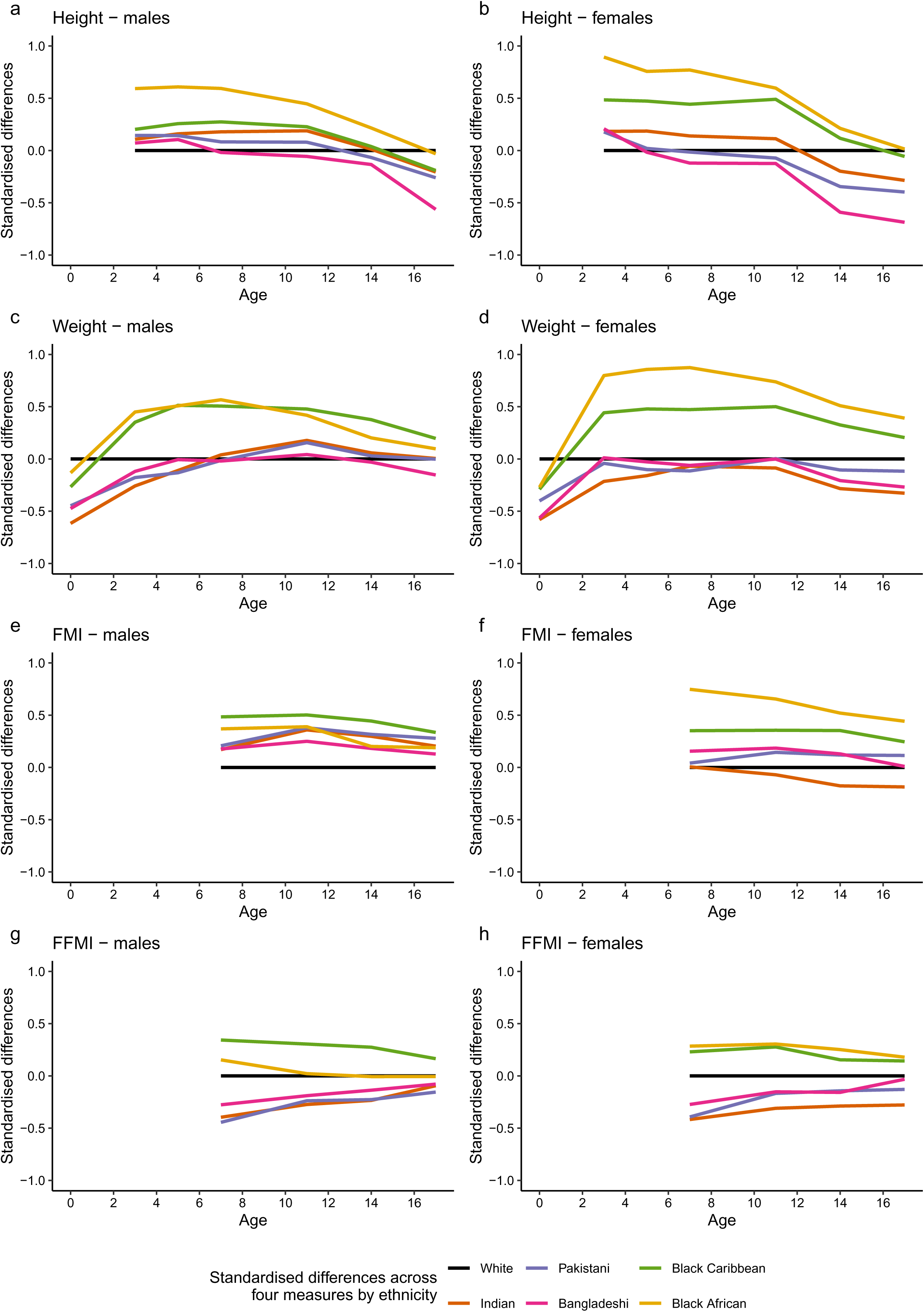
a-h: Standardised ethnic differences in height, weight, FMI, and FFMI compared to Whites stratified by sex

### Height from infancy through to age 17 by ethnicity

Height patterns were different. All minority boys and girls were taller than White children at age 3 (Figure 1, ESM Tables 2a and 2b). However, this height advantage was progressively lost in South Asian children: by age 7, Bangladeshi boys and girls were shorter than White children, by age 11, Pakistani children were shorter than White children, and finally by age 14, Indian children were shorter than White children. In boys and girls, by age 17, Indian children were 1.8/2.5 cm, Pakistani children 2.2/3.4 cm, and Bangladeshi children 4.8/6.0 cm shorter than White children (Table 2a). Black Caribbean and Black African children also displayed a progressive loss of height advantage with age, falling to similar or lower heights than White children only by age 17 (in boys and girls, 1.6/0.5 cm shorter in Black Caribbean children, 0.27 shorter in Black African boys, and only 0.1 cm taller in Black African girls compared to White children). The height gradient by ethnicity in girls aged 17 reflected that observed in their mothers.

**Table 2:** Accounting for ethnic differences (using white group as comparator) in FMI and FFMI at age 7 stratified by sex, coefficient and 95% CI.

|  |  | Boys |  |  |  |  | Girls |  |  |  |  |
| --- | --- | --- | --- | --- | --- | --- | --- | --- | --- | --- | --- |
|  |  | Indian | Pakistani | Bangladeshi | Black Caribbean | Black African | Indian | Pakistani | Bangladeshi | Black Caribbean | Black African |
| Fat Mass Index | Model 1 | 0.24<br>(-0.04, 0.53) | 0.29*<br>(0.06, 0.53) | 0.25<br>(-0.02, 0.52) | 0.68*<br>(0.16, 1.21) | 0.52***<br>(0.27, 0.78) | 0.01<br>(-0.24, 0.26) | 0.07<br>(-0.13, 0.27) | 0.26<br>(-0.11, 0.62) | 0.59**<br>(0.16, 1.01) | 1.24***<br>(0.81, 1.68) |
|  | Model 2 | 0.14<br>(-0.14, 0.43) | 0.24*<br>(0.01, 0.47) | 0.19<br>(-0.08, 0.46) | 0.58*<br>(0.07, 1.08) | 0.45***<br>(0.20, 0.70) | -0.09<br>(-0.36, 0.18) | -0.02<br>(-0.22, 0.18) | 0.10<br>(-0.27, 0.46) | 0.40<br>(-0.02, 0.82) | 1.06***<br>(0.65, 1.48) |
|  | Model 3 | 0.34*<br>(0.08, 0.60) | 0.27*<br>(0.05, 0.49) | 0.17<br>(-0.08, 0.43) | 0.46<br>(-0.01, 0.94) | 0.31**<br>(0.08, 0.54) | 0.20<br>(-0.07, 0.48) | -0.02<br>(-0.21, 0.17) | 0.02<br>(-0.33, 0.36) | 0.09<br>(-0.34, 0.51) | 0.72***<br>(0.33, 1.11) |
| Fat-Free Mass Index | Model 1 | -0.47***<br>(-0.71, -0.23) | -0.53***<br>(-0.70, -0.36) | -0.33*<br>(-0.60, -0.05) | 0.41*<br>(0.09, 0.73) | 0.18<br>(-0.07, 0.44) | -0.48***<br>(-0.68, -0.28) | -0.45***<br>(-0.59, -0.31) | -0.31*<br>(-0.56, -0.06) | 0.26*<br>(0.01, 0.52) | 0.33**<br>(0.08, 0.57) |
|  | Model 2 | -0.51***<br>(-0.75, -0.28) | -0.55***<br>(-0.72, -0.38) | -0.36*<br>(-0.63, -0.08) | 0.36*<br>(0.04, 0.68) | 0.15<br>(-0.10, 0.40) | -0.52***<br>(-0.72, -0.31) | -0.49***<br>(-0.62, -0.35) | -0.38**<br>(-0.63, -0.12) | 0.19<br>(-0.06, 0.44) | 0.26*<br>(0.02, 0.49) |
|  | Model 3 | -0.29**<br>(-0.51, -0.08) | -0.44***<br>(-0.60, -0.28) | -0.27<br>(-0.55, 0.01) | 0.25<br>(-0.04, 0.54) | 0.03<br>(-0.19, 0.26) | -0.32**<br>(-0.53, -0.10) | -0.45***<br>(-0.58, -0.31) | -0.40**<br>(-0.65, -0.15) | 0.00***<br>(1.00, -0.25) | 0.05<br>(0.65, -0.17) |

(\* p < 0.05, \*\* p < 0.01, \*\*\* p < 0.001)
Model 1: Unadjusted
Model 2: Adjusted for weight gain from birth to age 3
Model 3: Adjusted for maternal (height, pre-pregnancy weight, BMI, smoking, breastfeeding to 4 months), birthweight, weight gain from birth to age 3 and socioeconomic factors (index of multiple deprivation, parental education, parental income).

### Waist circumference by ethnicity

There were no clear differences in waist circumference and waist-to-height ratios by ethnicity at ages 5 and 7 years (Table 1).

### Ethnic differences in menstruation onset by age 11

Ethnic minority girls were 1.3-3 times more likely to have commenced menstruation by age 11 years than White girls (table 1).

### Ethnic differences in fat mass index and fat free mass index in childhood and adolescence

FMI was higher in all minority groups, and in particular in Black African and Black Caribbean boys and girls at ages 7, 11, 14, and 17. The exception was Indian girls who displayed lower FMI than their White counterparts throughout (Figure 1, ESM Tables 2a and 2b). FFMI was lower in all South Asian groups for both boys and girls, and lower in Black African boys than their White counterparts after age 7. Black Caribbean boys and girls had higher FFMI than White children at all ages. Ethnic differences in FFMI were most marked at age 7, with a progressive convergence to levels in White children by age 17 (Figure 1). The FMI/FFMI ratio at age 7 was elevated in all ethnic minority groups for both boys and girls compared to whites (table 1).

Adjustment for relative weight change from birth to age 3 years markedly attenuated ethnic differences in FMI at age 7 for both boys and girls (Table 2). Multivariable models indicated an additional contribution from maternal and socioeconomic factors for Black Caribbean and Black African children, but not South Asian children (Table 2, and ESM tables 3a and 3b). Similar, though less marked, patterns of attenuation of ethnic differences compared to White children were observed for FFMI (Table 2, and ESM tables 4a and 4b).

## Discussion

Young adults of South Asian, Black African and Black Caribbean ethnicity born in the UK at the turn of this century have distinct early life growth patterns compared to the White population. They share low birthweight and early accelerated growth, accompanied by premature growth deceleration. The magnitude of the former, and timing of the latter however differs by ethnicity, impacting final body habitus. At age 17, South Asian people were lighter and shorter, while Black African and Black Caribbean people were heavier, and of similar or lower height compared to their White counterparts. Minority ethnic groups had greater total fat mass than White people; but South Asian people had lower, and Black Caribbean children and Black African girls higher indices of fat free mass than White children. However, the ratio of fat to fat free mass was uniformly greater in all minority groups at age 7 than White children. Low birthweight followed by greater post-natal growth in all minority groups studied here contributes to excess fat mass index compared to White children, with further contributions in people of Black African and Black Caribbean ethnicity from adverse both individual and neighbourhood socioeconomic position.

Low birthweight in children of South Asian, Black Caribbean and Black African ethnicity in the UK is known^16^, and previous analyses of this cohort concluded that this was largely due to adverse socioeconomic circumstances^10^. While accelerated post-natal growth is also recognized^17^, the current analysis is the first to identify important differences in the effects this has on achieved height and weight beyond age 5 for different ethnicities. Relative weight gain from birth in all ethnic minority groups is of similar magnitude, and greater than that observed in White children. But as children of Black African and Black Caribbean ethnicity have higher birthweights than South Asian children, this accelerated weight gain results in heavier children by age 3 compared to White children, an advantage that persists through to late adolescence. Absolute weight gain in South Asian children is lower, and weight only briefly achieves levels similar to or greater than White boys around age 11, otherwise remaining lower than White boys and girls throughout early life.

In contrast to weight, all ethnic minority groups are taller by the age of 3 years. This height advantage is also progressively and differentially lost. By age 17, Black African children are of similar, and Black Caribbean children somewhat shorter than White children. The height advantage at age 3 in South Asian groups is progressively lost, firstly in those of Bangladeshi, then of Pakistani and finally of Indian origin. By age 14 all South Asian ethnicities are shorter than White children. This ethnic difference is particularly marked for Bangladeshi children, who by age 17 are ∼5.0-6.0 cm shorter than their White counterparts.

Interestingly the ethnic subgroup gradient in achieved height and weight by age 17 in the current generation is identical to that observed in their parents. Persistence of low birthweight has been observed some generations after migration from Surinam to the Netherlands^18^, and in African Americans in the US^19^, resident in the US for many generations. Short maternal stature and lower lean mass correlate with smaller organ mass and smaller birth canal dimensions^20,21^, which may then constrain fetal growth through both genetic and plastic mechanisms^22^. More broadly, poor in-utero development as a consequence of constrained early growth of the mother, and continuing adverse maternal circumstances may determine persistence of adverse growth patterns in migrant populations^20–22^. It should be noted that height is strongly influenced by environmental factors; for example, in just a few decades, South Koreans have become on average 8 cm taller than their North Korean counterparts.

Continued low socioeconomic position in minority groups is however a unifying feature in high income countries, and others have shown that low socioeconomic position is a key determinant of these low birthweights^10^. Most previous studies focus on the period of early weight gain^23^; but we show that while relative weight gain is similar across minority groups (and uniformly greater than in Whites) the absolute consequences for height and weight, allied to differential ‘wearing off’ of this rapid growth, is markedly different. Accelerated post-natal growth was observed in all minority groups, even though certain standard risk factors, such as maternal smoking, breastfeeding and maternal educational status were often more favourable.

All minorities displayed earlier growth deceleration compared to White children, and all minority girls were markedly more likely to experience early puberty, defined here as menstruation onset before age 11, than White girls. This confirms previous analyses on this cohort, but notably the percentage of Black Caribbean girls, at ∼18%, is far higher than previously reported (13.0%)^13^, as we excluded those of mixed ethnicity from our analysis. This earlier onset of menarche will make an additional contribution to adult obesity and type 2 diabetes mellitus^24^.

We confirm fat mass index at age 7 is greater in all minority groups^25^, and remains so through to age 17 apart from Indian girls. We also confirm that fat free mass index is lower in South Asian children, but higher in children of Black Caribbean, and girls of Black African ethnicity^6^. However, in resolution of this paradox in fat free mass, we also show that the ratio of fat-to-fat free mass is uniformly greater in all ethnic minority groups studied here compared to White children at age 7. Skeletal muscle, proxied here by fat free mass, is an important determinant of type 2 diabetes mellitus risk^26^. Mechanisms include impaired insulin stimulated glucose uptake, fatty deposition, imbalanced protein synthesis, and poor mitochondrial function. While bioelectrical impendence analysis captures quantity, and we show that relative quantity is lower in minority groups than in Whites, this tells us little about relative quality. Given the more adverse early life patterns of growth in minority groups, we suggest that lean mass quality may also be poorer^27^, and contribute to the observed greater risk and earlier onset of type 2 diabetes mellitus.

Biological, specifically genetic explanations have been invoked to account for ethnic differences in body composition^28^. However, self-reported ethnicity is not a biological construct^29^and the importance of current lifestyle, and of socioeconomic position should not be overlooked.

Generational increases in height^8^, by some 6 cm over 50 years in the UK, have been ascribed to better diets and to a reduction in catastrophic childhood infection. Socioeconomic status, as a determinant of access to both adequate and healthy nutrition may account for the reversal in the socioeconomic gradient in obesity^8^, and for the more adverse fat patterning observed in latter generations.

Ethnic differences in fat mass index were most strongly associated with relative weight gain between birth and age 3. Birthweight and maternal pre-pregnancy weight also made independent contributions to ethnic differences in fat mass index. It should be noted that South Asian mothers were both lighter and shorter than their White counterparts, which accounts for the multivariable estimated ethnic difference in fat mass index in South Asian versus White children being of greater magnitude than the unadjusted estimate, as generally lower maternal height and weight should each associate with lower rather than higher fat mass index in the offspring. While we distinguish maternal/early life and socioeconomic determinants of ethnic differences in fat mass, it should be noted that the former, such as maternal height, smoking status, and offspring birthweight, are also dependent on socioeconomic factors, thus it is likely that we have underestimated the role of socioeconomic position in accounting for ethnic differences. Additionally, similar to previous analyses of socioeconomic differentials^9^, we show that individual and neighbourhood socioeconomic position each contribute to ethnic differences in fat mass, indicating that beyond familial circumstances, the more obesogenic environments of neighbourhood deprivation (greater density of fast food outlets, fewer opportunities for recreation), play a role in adverse fat accumulation. Yet adverse neighbourhood socioeconomic factors, which should similarly affect people of both South Asian and of African ethnicity, cannot explain the markedly lower levels of fat free mass in South Asian groups. Pomeroy and colleagues demonstrate a marked reduction in stature in people from the Indian subcontinent allied to adoption of an agricultural lifestyle some 11,000 years ago, and of a low lean mass for given body size^30^. They hypothesise that these expressions of body habitus are due to historical climatic adaptation and to cumulative ecological pressures.

The MCS is representative of the UK population at birth, and efforts were made to ensure appropriate representation of ethnic minority groups. Sample attrition, and attendant data missingness, has been overcome by multiple imputation. We largely report absolute (standardised) differences in body habitus at each age, rather than model growth trajectories. This is a critical initial first step to identify important differences and critical time points before embarking on methods to model growth trajectory heterogeneity, which often make a priori assumptions, require careful specification, and ideally multiple approaches to compare consistency of outcomes and thus enable robustness of conclusions^31^. Bioelectrical impedence analysis estimate fat mass% and can be used to derive fat mass and fat free mass, and the respective indexes. These equations make assumptions about relationships between body habitus and fat mass, and about the relationship of body proportions with fat-free mass. Previous BIA validation studies against gold standard deuterium methods in UK multi-ethnic childhood and adolescent populations variously report both over and underestimation of fat mass and fat free mass by bioimpedance in ethnic minority groups^6,32^. Bioimpedance equations which overcome these biases have been proposed. Unfortunately, the MCS did not record direct bioimpedance measures, so that more ethnically valid derived equations could not be applied. However, as the direction of ethnic difference in fat mass and fat free mass indices remains consistent in validation studies, even though the magnitude may differ^6^ we do not view this as a major limitation.

In conclusion, we demonstrate marked ethnic subgroup differences in early development that relate to achieved height and weight, body fat, fat free mass and reproductive maturity in the UK. Ethnic minority groups in the UK display more adverse patterns in body habitus than White children, likely determining early onset of type 2 diabetes mellitus and other chronic conditions, though further follow up of MCS is needed to be certain of this. The role of ethnic differences in quality, as opposed to quantity of fat and of lean mass in accounting for differential risk of type 2 diabetes mellitus is not known, nor is the impact of disordered early growth on tissue quality understood. Part of these ethnic differences can be accounted for by adverse individual and neighbourhood socioeconomic position, which have persisted across generations^33^. Addressing socioeconomic adversity is imperative if we are to resolve the inter-generational cycle of adverse growth and premature onset of chronic disease.

## Data Availability

Data are freely available to bona fide researchers under standard access conditions via the UK Data Service
http://ukdataservice.ac.uk

## Acknowledgements

We would like to thank all families participating in the Millennium Cohort Study.

## Authors’ relationships and activities

NC reports receiving funds from AstraZeneca for serving on data safety and management committees of clinical trials sponsored by the company. The remaining authors have no relationships to declare

## Contribution Statement

NC conceived the broad idea for the analysis. NC, KP, SE and CB-S designed the study and analysis plan, reviewed and interpreted tables, and drafted the paper. KP performed the statistical analysis. All authors contributed to further review and redraft of the paper. All authors approved the final version and decision to submit. NC is responsible for the integrity of the work as a whole.

## Data availability

All MCS data used in this analysis are available from UK Data Service with an end user license. https://ukdataservice.ac.uk/find-data/

## Abbreviations

FMI: fat mass index
FFMI: fat free mass index
MCS: Millennium Cohort Study

