## Supplemental tables for "Diabetogenic elevated childhood total fat in South Asian and Black African/Caribbean people relates to adverse early life growth and low socioeconomic position compared to White people in the UK"

| **ESM Table 1. Numbers for missing data by variable, subsequently imputed** | | | | |
| --- | --- | --- | --- | --- |
|  | **Variable** | | **Not Missing** | **Missing** |
|  | **Birth:** | Weight | 13,980 | 13 |
|  |  | Maternal Height | 13,814 | 179 |
|  |  | Maternal Weight | 12,965 | 1,028 |
|  |  | Maternal Smoking | 9,356 | 4,637 |
|  |  | Maternal Breastfeeding | 9,336 | 4,657 |
|  |  | Area Deprivation | 13,992 | 1 |
|  |  | Highest Parental Income | 13,964 | 29 |
|  |  | Highest Parental Education | 13,961 | 32 |
|  | **Age 3:** | Height | 11,633 | 2,360 |
|  |  | Weight | 11,781 | 2,212 |
|  | **Age 5:** | Height | 13,502 | 491 |
|  |  | Weight | 13,522 | 471 |
|  |  | Waist | 13,420 | 573 |
|  | **Age 7:** | Height | 11,832 | 2,161 |
|  |  | Weight | 11,803 | 2,190 |
|  |  | Waist | 11,722 | 2,271 |
|  |  | Fat Mass | 11,632 | 2,361 |
|  | **Age 11:** | Height | 11,087 | 2,906 |
|  |  | Weight | 10,909 | 3,084 |
|  |  | Fat Mass | 10,785 | 3,208 |
|  | **Age 14:** | Height | 9,604 | 4,389 |
|  |  | Weight | 9,347 | 4,646 |
|  |  | Fat Mass | 9,225 | 4,768 |
|  | **Age 17:** | Height | 8,205 | 5,788 |
|  |  | Weight | 7,996 | 5,997 |
|  |  | Fat Mass | 7,875 | 6,118 |

| Age | | **Boys** | | | | | | **Girls** | | | | | |
| --- | --- | --- | --- | --- | --- | --- | --- | --- | --- | --- | --- | --- | --- |
|  |  | **White** | **Indian** | **Pakistani** | **Bangladeshi** | **Black Caribbean** | **Black African** | **White** | **Indian** | **Pakistani** | **Bangladeshi** | **Black Caribbean** | **Black African** |
| **Height (cm)** | **3** | 95.93 ± 4.65 | 96.44 ± 4.84 | 96.62 ± 4.49 | 96.27 ± 6.51 | 96.89 ± 5.26 | 98.74 ± 5.77 | 94.74 ± 4.55 | 95.57 ± 6.12 | 95.54 ± 5.14 | 95.68 ± 4.75 | 96.94 ± 4.33 | 98.80 ± 5.76 |
|  | **5** | 111.08 ± 5.46 | 111.98 ± 6.08 | 111.88 ± 5.67 | 111.67 ± 6.75 | 112.52 ± 6.79 | 114.48 ± 6.96 | 110.18 ± 5.51 | 111.21 ± 6.89 | 110.29 ± 5.62 | 110.09 ± 6.23 | 112.79 ± 4.85 | 114.35 ± 6.06 |
|  | **7** | 124.07 ± 6.24 | 125.21 ± 6.75 | 124.60 ± 6.36 | 123.95 ± 8.36 | 125.82 ± 7.17 | 127.85 ± 7.96 | 123.17 ± 6.30 | 124.05 ± 7.62 | 123.08 ± 6.12 | 122.42 ± 6.74 | 125.96 ± 4.77 | 128.03 ± 7.44 |
|  | **11** | 145.98 ± 8.11 | 147.54 ± 9.46 | 146.64 ± 8.07 | 145.51 ± 8.97 | 147.86 ± 9.31 | 149.68 ± 9.83 | 146.83 ± 8.58 | 147.79 ± 10.43 | 146.21 ± 8.39 | 145.76 ± 8.99 | 151.04 ± 7.47 | 151.95 ± 9.71 |
|  | **14** | 167.12 ± 9.66 | 167.24 ± 9.30 | 166.45 ± 8.80 | 165.79 ± 10.50 | 167.49 ± 10.65 | 169.24 ± 10.75 | 161.82 ± 8.20 | 160.20 ± 9.50 | 159.00 ± 7.39 | 156.98 ± 8.60 | 162.78 ± 7.59 | 163.56 ± 9.07 |
|  | **17** | 177.04 ± 8.39 | 175.27 ± 8.17 | 174.80 ± 7.03 | 172.21 ± 9.96 | 175.40 ± 9.25 | 176.77 ± 9.51 | 164.36 ± 8.66 | 161.88 ± 9.41 | 160.92 ± 6.78 | 158.41 ± 8.20 | 163.87 ± 7.36 | 164.48 ± 8.21 |
| **Weight (Kg)** | **0** | 3.46 ± 0.68 | 3.03 ± 0.89 | 3.15 ± 0.65 | 3.13 ± 0.61 | 3.27 ± 0.87 | 3.37 ± 0.73 | 3.34 ± 0.66 | 2.96 ± 0.81 | 3.08 ± 0.67 | 2.97 ± 0.95 | 3.15 ± 0.72 | 3.16 ± 0.72 |
|  | **3** | 15.30 ± 2.26 | 14.70 ± 2.84 | 14.89 ± 2.56 | 15.02 ± 2.92 | 16.11 ± 2.90 | 16.33 ± 3.00 | 14.63 ± 2.27 | 14.15 ± 2.98 | 14.54 ± 2.62 | 14.66 ± 3.49 | 15.64 ± 2.92 | 16.45 ± 3.70 |
|  | **5** | 20.23 ± 3.12 | 19.88 ± 4.24 | 19.81 ± 4.05 | 20.21 ± 4.51 | 21.87 ± 4.46 | 21.85 ± 4.30 | 19.80 ± 3.31 | 19.27 ± 3.68 | 19.46 ± 3.73 | 19.70 ± 4.80 | 21.38 ± 3.73 | 22.63 ± 4.65 |
|  | **7** | 25.50 ± 4.91 | 25.70 ± 6.68 | 25.43 ± 6.80 | 25.40 ± 6.48 | 28.05 ± 7.00 | 28.35 ± 6.78 | 25.33 ± 5.33 | 24.97 ± 5.64 | 24.72 ± 5.54 | 25.00 ± 6.83 | 27.85 ± 5.67 | 29.99 ± 7.95 |
|  | **11** | 40.51 ± 10.66 | 42.44 ± 13.69 | 42.20 ± 12.59 | 40.97 ± 12.63 | 45.71 ± 14.97 | 45.06 ± 13.12 | 42.07 ± 11.62 | 41.07 ± 12.15 | 42.11 ± 12.27 | 42.06 ± 12.67 | 47.89 ± 12.91 | 50.64 ± 14.80 |
|  | **14** | 58.57 ± 15.12 | 59.48 ± 18.33 | 59.03 ± 16.40 | 58.10 ± 15.59 | 64.39 ± 20.50 | 61.69 ± 16.73 | 57.81 ± 14.93 | 53.57 ± 13.64 | 56.23 ± 16.39 | 54.72 ± 15.45 | 62.67 ± 17.58 | 65.42 ± 18.54 |
|  | **17** | 71.87 ± 19.48 | 71.97 ± 21.59 | 71.84 ± 20.83 | 68.83 ± 20.02 | 75.79 ± 21.55 | 73.81 ± 19.86 | 64.02 ± 17.20 | 58.39 ± 17.12 | 62.01 ± 18.93 | 59.39 ± 18.34 | 67.54 ± 20.39 | 70.75 ± 20.62 |
| **FMI (kg/m^2^)** | **7** | 3.34 ± 1.38 | 3.58 ± 1.89 | 3.63 ± 2.13 | 3.59 ± 1.62 | 4.02 ± 2.36 | 3.86 ± 1.47 | 3.74 ± 1.66 | 3.75 ± 1.65 | 3.81 ± 1.80 | 4.00 ± 2.17 | 4.33 ± 1.94 | 4.99 ± 2.50 |
|  | **11** | 3.91 ± 2.52 | 4.83 ± 3.23 | 4.89 ± 3.09 | 4.55 ± 3.16 | 5.20 ± 4.63 | 4.91 ± 3.06 | 4.96 ± 2.92 | 4.75 ± 2.75 | 5.38 ± 3.08 | 5.50 ± 3.25 | 6.00 ± 3.54 | 6.87 ± 3.69 |
|  | **14** | 3.66 ± 2.76 | 4.50 ± 3.74 | 4.55 ± 3.38 | 4.17 ± 2.95 | 4.91 ± 4.49 | 4.22 ± 2.97 | 6.24 ± 3.52 | 5.62 ± 3.00 | 6.65 ± 4.02 | 6.69 ± 3.85 | 7.48 ± 4.35 | 8.07 ± 4.38 |
|  | **17** | 3.94 ± 3.53 | 4.68 ± 4.44 | 4.95 ± 4.30 | 4.41 ± 3.67 | 5.15 ± 4.76 | 4.63 ± 3.55 | 7.05 ± 4.43 | 6.23 ± 3.92 | 7.56 ± 5.02 | 7.10 ± 4.80 | 8.14 ± 5.73 | 9.01 ± 5.13 |
| **FFMI (kg/m^2^)** | **7** | 13.16 ± 1.16 | 12.69 ± 1.57 | 12.63 ± 1.51 | 12.83 ± 1.65 | 13.56 ± 1.45 | 13.34 ± 1.46 | 12.87 ± 1.15 | 12.39 ± 1.32 | 12.42 ± 1.24 | 12.56 ± 1.50 | 13.14 ± 1.13 | 13.20 ± 1.41 |
|  | **11** | 14.97 ± 1.78 | 14.47 ± 2.08 | 14.54 ± 1.88 | 14.63 ± 2.13 | 15.53 ± 1.95 | 15.01 ± 2.03 | 14.41 ± 1.62 | 13.91 ± 1.96 | 14.14 ± 1.88 | 14.16 ± 1.93 | 14.86 ± 1.62 | 14.91 ± 1.70 |
|  | **14** | 17.21 ± 2.39 | 16.64 ± 2.47 | 16.66 ± 2.50 | 16.88 ± 2.57 | 17.88 ± 2.74 | 17.20 ± 2.63 | 15.78 ± 1.97 | 15.21 ± 2.20 | 15.50 ± 2.12 | 15.47 ± 2.26 | 16.08 ± 2.04 | 16.28 ± 1.85 |
|  | **17** | 18.96 ± 2.83 | 18.69 ± 2.49 | 18.51 ± 2.81 | 18.73 ± 2.73 | 19.43 ± 2.80 | 18.94 ± 3.40 | 16.63 ± 2.32 | 15.98 ± 2.48 | 16.33 ± 2.34 | 16.55 ± 2.88 | 16.96 ± 2.51 | 17.04 ± 2.32 |

**ESM Table 2a) Height, weight, fat mass index, and fat-free mass index (mean +/- standard deviation) by age, ethnicity, and sex**

**ESM Table 2b) Standardised height, weight, fat mass index, and fat-free mass index by age ethnicity, and sex**

| **Age** | | **Boys** | | | | | | **Girls** | | | | | |
| --- | --- | --- | --- | --- | --- | --- | --- | --- | --- | --- | --- | --- | --- |
|  |  | **White** | **Indian** | **Pakistani** | **Bangladeshi** | **Black Caribbean** | **Black African** | **White** | **Indian** | **Pakistani** | **Bangladeshi** | **Black Caribbean** | **Black African** |
| **Height (cm)** | **3** | - | 0.11 | 0.14 | 0.07 | 0.20 | 0.59 | - | 0.18 | 0.18 | 0.21 | 0.48 | 0.89 |
|  | **5** | - | 0.16 | 0.14 | 0.10 | 0.26 | 0.61 | - | 0.19 | 0.02 | -0.02 | 0.47 | 0.76 |
|  | **7** | - | 0.18 | 0.08 | -0.02 | 0.27 | 0.59 | - | 0.14 | -0.01 | -0.12 | 0.44 | 0.77 |
|  | **11** | - | 0.19 | 0.08 | -0.06 | 0.23 | 0.45 | - | 0.11 | -0.07 | -0.12 | 0.49 | 0.60 |
|  | **14** | - | 0.01 | -0.07 | -0.13 | 0.04 | 0.22 | - | -0.20 | -0.35 | -0.59 | 0.12 | 0.21 |
|  | **17** | - | -0.21 | -0.26 | -0.56 | -0.19 | -0.03 | - | -0.29 | -0.40 | -0.69 | -0.06 | 0.01 |
| **Weight (Kg)** | **0** | - | -0.62 | -0.45 | -0.47 | -0.27 | -0.13 | - | -0.58 | -0.40 | -0.56 | -0.29 | -0.27 |
|  | **3** | - | -0.26 | -0.18 | -0.12 | 0.35 | 0.45 | - | -0.21 | -0.04 | 0.01 | 0.44 | 0.80 |
|  | **5** | - | -0.11 | -0.13 | -0.01 | 0.51 | 0.51 | - | -0.16 | -0.10 | -0.03 | 0.48 | 0.86 |
|  | **7** | - | 0.04 | -0.01 | -0.02 | 0.51 | 0.57 | - | -0.07 | -0.11 | -0.06 | 0.47 | 0.87 |
|  | **11** | - | 0.18 | 0.16 | 0.04 | 0.48 | 0.42 | - | -0.09 | 0.00 | 0.00 | 0.50 | 0.74 |
|  | **14** | - | 0.06 | 0.03 | -0.03 | 0.38 | 0.20 | - | -0.28 | -0.11 | -0.21 | 0.33 | 0.51 |
|  | **17** | - | 0.00 | 0.00 | -0.15 | 0.20 | 0.10 | - | -0.33 | -0.12 | -0.27 | 0.20 | 0.39 |
| **FMI (kg/m^2^)** | **7** | - | 0.17 | 0.21 | 0.18 | 0.48 | 0.37 | - | 0.01 | 0.04 | 0.16 | 0.35 | 0.75 |
|  | **11** | - | 0.36 | 0.38 | 0.25 | 0.50 | 0.39 | - | -0.07 | 0.14 | 0.18 | 0.36 | 0.65 |
|  | **14** | - | 0.30 | 0.32 | 0.18 | 0.44 | 0.20 | - | -0.18 | 0.12 | 0.13 | 0.35 | 0.52 |
|  | **17** | - | 0.20 | 0.28 | 0.13 | 0.34 | 0.19 | - | -0.19 | 0.11 | 0.01 | 0.25 | 0.44 |
| **FFMI (kg/m^2^)** | **7** | - | -0.40 | -0.44 | -0.28 | 0.34 | 0.15 | - | -0.42 | -0.39 | -0.27 | 0.23 | 0.28 |
|  | **11** | - | -0.27 | -0.24 | -0.19 | 0.30 | 0.02 | - | -0.31 | -0.17 | -0.15 | 0.28 | 0.31 |
|  | **14** | - | -0.23 | -0.23 | -0.14 | 0.27 | -0.01 | - | -0.29 | -0.14 | -0.16 | 0.15 | 0.25 |
|  | **17** | - | -0.09 | -0.15 | -0.08 | 0.16 | -0.01 | - | -0.28 | -0.13 | -0.03 | 0.14 | 0.18 |

**ESM Table 3a.** **Accounting for ethnic differences (using white group as comparator) in FMI at age 7 in boys, coefficient and 95% CI, p value**

|  | **Indian** | | **Pakistani** | | **Bangladeshi** | | **Black Caribbean** | | **Black African** | |
| --- | --- | --- | --- | --- | --- | --- | --- | --- | --- | --- |
|  | *Coefficient (95% CI)* | *P value* | *Coefficient (95% CI)* | *P value* | *Coefficient (95% CI)* | *P value* | *Coefficient (95% CI)* | *P value* | *Coefficient (95% CI)* | *P value* |
| **Model 1** |  |  |  |  |  |  |  |  |  |  |
| Ethnicity | 0.24  (-0.04, 0.53) | 0.097 | 0.29  (0.06, 0.53) | 0.014 | 0.25  (-0.02, 0.52) | 0.071 | 0.68  (0.16, 1.21) | 0.01 | 0.52  (0.27, 0.78) | <0.001 |
| **Model 2** |  |  |  |  |  |  |  |  |  |  |
| Ethnicity | 0.14  (-0.14, 0.43) | 0.322 | 0.24  (0.01, 0.47) | 0.042 | 0.19  (-0.08, 0.46) | 0.175 | 0.58  (0.07, 1.08) | 0.026 | 0.45  (0.20, 0.70) | <0.001 |
| Weight gain to age 3 | 0.17  (0.13, 0.22) | <0.001 | 0.18  (0.13, 0.22) | <0.001 | 0.17  (0.12, 0.22) | <0.001 | 0.17  (0.13, 0.22) | <0.001 | 0.17  (0.13, 0.22) | <0.001 |
| **Model 3** |  |  |  |  |  |  |  |  |  |  |
| Ethnicity | 0.34  (0.08, 0.60) | 0.012 | 0.2  7(0.05, 0.49) | 0.017 | 0.17  (-0.09, 0.43) | 0.194 | 0.45  (-0.02, 0.93) | 0.062 | 0.31  (0.08, 0.54) | 0.009 |
| Weight gain to age 3 | 0.48  (0.39, 0.58) | <0.001 | 0.49  (0.40, 0.59) | <0.001 | 0.48  (0.39, 0.58) | <0.001 | 0.48  (0.39, 0.58) | <0.001 | 0.49  (0.39, 0.58) | <0.001 |
| Maternal height | -2.54  (-3.07, -2.02) | <0.001 | -2.58  (-3.10, -2.06) | <0.001 | -2.61  (-3.13, -2.09) | <0.001 | -2.62  (-3.14, -2.09) | <0.001 | -2.56  (-3.08, -2.04) | <0.001 |
| Maternal weight | 0.02  (0.02, 0.02) | <0.001 | 0.0  2(0.02, 0.02) | <0.001 | 0.02  (0.02, 0.02) | <0.001 | 0.02  (0.02, 0.02) | <0.001 | 0.02  (0.02, 0.02) | <0.001 |
| Breast fed to 4 months | -0.09  (-0.18, 0.00) | 0.061 | -0.08  (-0.18, 0.01) | 0.069 | -0.08  (-0.18, 0.01) | 0.086 | -0.08  (-0.17, 0.02) | 0.114 | -0.08  (-0.17, 0.02) | 0.101 |
| Maternal smoking | 0.10  (-0.03, 0.23) | 0.134 | 0.10  (-0.03, 0.23) | 0.126 | 0.10  (-0.03, 0.23) | 0.12 | 0.11  (-0.02, 0.24) | 0.108 | 0.10  (-0.03, 0.23) | 0.124 |
| Birthweight | 0.85  (0.73, 0.97) | <0.001 | 0.87  (0.74, 0.99) | <0.001 | 0.85  (0.73, 0.97) | <0.001 | 0.85  (0.73, 0.97) | <0.001 | 0.85  (0.73, 0.97) | <0.001 |
| Area deprivation | -0.02  (-0.03, 0.00) | 0.022 | -0.01  (-0.03, 0.00) | 0.058 | -0.01  (-0.03, 0.00) | 0.031 | -0.01  (-0.03, 0.00) | 0.032 | -0.02  (-0.03, 0.00) | 0.018 |
| Income | -0.05  (-0.08, -0.02) | <0.001 | -0.05  (-0.08, -0.02) | <0.001 | -0.05  (-0.08, -0.02) | <0.001 | -0.05  (-0.08, -0.02) | <0.001 | -0.05  (-0.07, -0.02) | 0.001 |

**ESM Table 3b. Accounting for ethnic differences (using white group as comparator) in FMI at age 7 in girls, coefficient and 95% CI, p value**

|  | **Indian** | | **Pakistani** | | **Bangladeshi** | | **Black Caribbean** | | **Black African** | |
| --- | --- | --- | --- | --- | --- | --- | --- | --- | --- | --- |
|  | Coefficient (95% CI) | P value | Coefficient (95% CI) | P value | Coefficient (95% CI) | P value | Coefficient (95% CI) | P value | Coefficient (95% CI) | P value |
| **Model 1** |  |  |  |  |  |  |  |  |  |  |
| Ethnicity | 0.01  (-0.24, 0.26) | 0.942 | 0.07  (-0.13, 0.27) | 0.503 | 0.26  (-0.11, 0.62) | 0.164 | 0.59  (0.16, 1.01) | 0.008 | 1.24  (0.81, 1.68) | <0.001 |
| **Model 2** |  |  |  |  |  |  |  |  |  |  |
| Ethnicity | -0.09  (-0.36, 0.18) | 0.516 | -0.02  (-0.22, 0.18) | 0.851 | 0.10  (-0.27, 0.46) | 0.601 | 0.40  (-0.02, 0.82) | 0.059 | 1.06  (0.65, 1.48) | <0.001 |
| Weight gain to age 3 | 0.20  (0.15, 0.26) | <0.001 | 0.21  (0.15, 0.26) | <0.001 | 0.21  (0.15, 0.27) | <0.001 | 0.21  (0.15, 0.26) | <0.001 | 0.21  (0.16, 0.27) | <0.001 |
| **Model 3** |  |  |  |  |  |  |  |  |  |  |
| Ethnicity | 0.20  (-0.08, 0.47) | 0.158 | -0.02  (-0.20, 0.17) | 0.847 | 0.02  (-0.32, 0.37) | 0.901 | 0.09  (-0.34, 0.52) | 0.683 | 0.72  (0.33, 1.11) | <0.001 |
| Weight gain to age 3 | 0.61  (0.48, 0.73) | <0.001 | 0.60  (0.47, 0.72) | <0.001 | 0.61  (0.49, 0.74) | <0.001 | 0.59  (0.47, 0.71) | <0.001 | 0.62  (0.49, 0.75) | <0.001 |
| Maternal height | -2.26  (-2.91, -1.60) | <0.001 | -2.21  (-2.86, -1.56) | <0.001 | -2.24  (-2.90, -1.57) | <0.001 | -2.19  (-2.85, -1.53) | <0.001 | -2.14  (-2.80, -1.49) | <0.001 |
| Maternal weight | 0.02  (0.02, 0.02) | <0.001 | 0.02  (0.02, 0.03) | <0.001 | 0.02  (0.02, 0.02) | <0.001 | 0.02  (0.02, 0.03) | <0.001 | 0.02  (0.02, 0.02) | <0.001 |
| Breast fed to 4 months | -0.12  (-0.21, -0.03) | 0.012 | -0.11  (-0.21, -0.02) | 0.018 | -0.11(-0.20, -0.01) | 0.025 | -0.11  (-0.21, -0.02) | 0.023 | -0.11  (-0.21, -0.02) | 0.021 |
| Maternal smoking | 0.17  (0.03, 0.30) | 0.017 | 0.17  (0.04, 0.31) | 0.012 | 0.18  (0.04, 0.31) | 0.01 | 0.17  (0.04, 0.31) | 0.013 | 0.18  (0.04, 0.31) | 0.01 |
| Birthweight | 1.19  (1.04, 1.35) | <0.001 | 1.17  (1.01, 1.33) | <0.001 | 1.20  (1.04, 1.36) | <0.001 | 1.16  (1.01, 1.32) | <0.001 | 1.20  (1.04, 1.36) | <0.001 |
| Area deprivation | -0.02  (-0.03, -0.01) | 0.008 | -0.02  (-0.04, -0.01) | 0.004 | -0.02  (-0.04, -0.01) | 0.005 | -0.02  (-0.04, -0.01) | 0.006 | -0.02  (-0.03, 0.00) | 0.011 |
| Income | -0.08  (-0.11, -0.04) | <0.001 | -0.07  (-0.10, -0.04) | <0.001 | -0.08  (-0.11, -0.04) | <0.001 | -0.07  (-0.11, -0.04) | <0.001 | -0.08  (-0.11, -0.04) | <0.001 |

**ESM Table 4a. Accounting for ethnic differences (using white group as comparator) in FFMI at age 7 in boys, coefficient and 95% CI, p value**

|  | **Indian** | | **Pakistani** | | **Bangladeshi** | | **Black Caribbean** | | **Black African** | |
| --- | --- | --- | --- | --- | --- | --- | --- | --- | --- | --- |
|  | Coefficient (95% CI) | P value | Coefficient (95% CI) | P value | Coefficient (95% CI) | P value | Coefficient (95% CI) | P value | Coefficient (95% CI) | P value |
| **Model 1** |  |  |  |  |  |  |  |  |  |  |
| Ethnicity | -0.47  (-0.71, -0.23) | <0.001 | -0.53  (-0.70, -0.36) | <0.001 | -0.33  (-0.60, -0.05) | 0.019 | 0.41  (0.09, 0.73) | 0.013 | 0.18  (-0.07, 0.44) | 0.159 |
| **Model 2** |  |  |  |  |  |  |  |  |  |  |
| Ethnicity | -0.51  (-0.75, -0.28) | <0.001 | -0.55  (-0.72, -0.38) | <0.001 | -0.36  (-0.63, -0.08) | 0.011 | 0.36  (0.04, 0.68) | 0.03 | 0.15  (-0.10, 0.40) | 0.247 |
| Weight gain to age 3 | 0.08  (0.04, 0.12) | <0.001 | 0.08  (0.04, 0.12) | <0.001 | 0.08  (0.04, 0.12) | <0.001 | 0.08  (0.04, 0.12) | <0.001 | 0.08  (0.04, 0.12) | <0.001 |
| **Model 3** |  |  |  |  |  |  |  |  |  |  |
| Ethnicity | -0.30  (-0.51, -0.08) | 0.007 | -0.44  (-0.60, -0.29) | <0.001 | -0.28  (-0.56, 0.00) | 0.054 | 0.25  (-0.03, 0.54) | 0.084 | 0.03  (-0.19, 0.26) | 0.788 |
| Weight gain to age 3 | 0.42  (0.32, 0.51) | <0.001 | 0.42  (0.33, 0.52) | <0.001 | 0.42  (0.32, 0.51) | <0.001 | 0.41  (0.32, 0.51) | <0.001 | 0.42  (0.32, 0.51) | <0.001 |
| Maternal height | -1.73  (-2.18, -1.28) | <0.001 | -1.74  (-2.19, -1.29) | <0.001 | -1.75  (-2.20, -1.30) | <0.001 | -1.75  (-2.20, -1.30) | <0.001 | -1.73  (-2.18, -1.28) | <0.001 |
| Maternal weight | 0.01  (0.01, 0.02) | <0.001 | 0.01  (0.01, 0.02) | <0.001 | 0.01  (0.01, 0.02) | <0.001 | 0.01  (0.01, 0.02) | <0.001 | 0.01  (0.01, 0.02) | <0.001 |
| Breast fed to 4 months | 0.00  (-0.07, 0.07) | 0.971 | 0.00  (-0.07, 0.08) | 0.944 | 0.00  (-0.07, 0.08) | 0.92 | 0.01  (-0.07, 0.08) | 0.873 | 0.00  (-0.07, 0.08) | 0.942 |
| Maternal smoking | 0.08  (-0.02, 0.19) | 0.104 | 0.09  (-0.01, 0.20) | 0.083 | 0.08  (-0.02, 0.19) | 0.108 | 0.09  (-0.02, 0.19) | 0.098 | 0.08  (-0.02, 0.19) | 0.106 |
| Birthweight | 0.91  (0.80, 1.02) | <0.001 | 0.91  (0.80, 1.02) | <0.001 | 0.90  (0.79, 1.01) | <0.001 | 0.91  (0.79, 1.02) | <0.001 | 0.91  (0.80, 1.02) | <0.001 |
| Area deprivation | 0.00  (-0.02, 0.01) | 0.364 | 0.00  (-0.01, 0.01) | 0.422 | 0.00  (-0.01, 0.01) | 0.401 | 0.00  (-0.01, 0.01) | 0.436 | 0.00  (-0.01, 0.01) | 0.408 |
| Income | -0.04  (-0.06, -0.01) | 0.002 | -0.04  (-0.06, -0.01) | 0.003 | -0.04  (-0.06, -0.01) | 0.003 | -0.04  (-0.06, -0.01) | 0.003 | -0.03  (-0.06, -0.01) | 0.005 |

**ESM Table 4b. Accounting for ethnic differences (using white group as comparator) in FFMI at age 7 in girls, coefficient and 95% CI, p value**

|  | **Indian** | | **Pakistani** | | **Bangladeshi** | | **Black Caribbean** | | **Black African** | |
| --- | --- | --- | --- | --- | --- | --- | --- | --- | --- | --- |
|  | Coefficient (95% CI) | P value | Coefficient (95% CI) | P value | Coefficient (95% CI) | P value | Coefficient (95% CI) | P value | Coefficient (95% CI) | P value |
| **Model 1** |  |  |  |  |  |  |  |  |  |  |
| Ethnicity | -0.48  (-0.68, -0.28) | <0.001 | -0.45  (-0.59, -0.31) | <0.001 | -0.31  (-0.56, -0.06) | 0.014 | 0.26  (0.01, 0.52) | 0.039 | 0.33  (0.08, 0.57) | 0.009 |
| **Model 2** |  |  |  |  |  |  |  |  |  |  |
| Ethnicity | -0.52  (-0.72, -0.31) | <0.001 | -0.49  (-0.62, -0.35) | <0.001 | -0.38  (-0.63, -0.12) | 0.004 | 0.19  (-0.06, 0.44) | 0.129 | 0.26  (0.02, 0.49) | 0.036 |
| Weight gain to age 3 | 0.08  (0.04, 0.11) | <0.001 | 0.08  (0.05, 0.12) | <0.001 | 0.08  (0.05, 0.12) | <0.001 | 0.08  (0.05, 0.12) | <0.001 | 0.08  (0.05, 0.12) | <0.001 |
| **Model 3** |  |  |  |  |  |  |  |  |  |  |
| Ethnicity | -0.32  (-0.54, -0.11) | 0.004 | -0.44  (-0.57, -0.31) | <0.001 | -0.39  (-0.64, -0.14) | 0.002 | 0.00  (-0.25, 0.25) | 0.99 | 0.05  (-0.17, 0.28) | 0.641 |
| Weight gain to age 3 | 0.39  (0.30, 0.47) | <0.001 | 0.38  (0.30, 0.47) | <0.001 | 0.40  (0.31, 0.48) | <0.001 | 0.38  (0.30, 0.46) | <0.001 | 0.40  (0.31, 0.49) | <0.001 |
| Maternal height | -1.60  (-2.08, -1.13) | <0.001 | -1.56  (-2.03, -1.09) | <0.001 | -1.58  (-2.05, -1.10) | <0.001 | -1.55  (-2.03, -1.08) | <0.001 | -1.56  (-2.03, -1.09) | <0.001 |
| Maternal weight | 0.01  (0.01, 0.01) | <0.001 | 0.01  (0.01, 0.01) | <0.001 | 0.01  (0.01, 0.01) | <0.001 | 0.01  (0.01, 0.01) | <0.001 | 0.01  (0.01, 0.01) | <0.001 |
| Breast fed to 4 months | -0.02  (-0.09, 0.06) | 0.633 | -0.01  (-0.08, 0.06) | 0.796 | -0.01  (-0.08, 0.07) | 0.829 | -0.01  (-0.09, 0.07) | 0.786 | -0.01  (-0.09, 0.06) | 0.741 |
| Maternal smoking | 0.10  (0.01, 0.19) | 0.028 | 0.11  (0.02, 0.20) | 0.017 | 0.11  (0.02, 0.20) | 0.015 | 0.11  (0.02, 0.19) | 0.018 | 0.11  (0.02, 0.20) | 0.016 |
| Birthweight | 0.92  (0.81, 1.02) | <0.001 | 0.90  (0.79, 1.01) | <0.001 | 0.92  (0.82, 1.03) | <0.001 | 0.90  (0.79, 1.00) | <0.001 | 0.92  (0.82, 1.03) | <0.001 |
| Area deprivation | -0.01  (-0.02, 0.00) | 0.088 | -0.01  (-0.02, 0.00) | 0.06 | -0.01  (-0.02, 0.00) | 0.069 | -0.01  (-0.02, 0.00) | 0.08 | -0.01  (-0.02, 0.00) | 0.09 |
| Income | -0.03  (-0.06, -0.01) | 0.004 | -0.03  (-0.05, -0.01) | 0.008 | -0.03  (-0.05, -0.01) | 0.005 | -0.03  (-0.05, -0.01) | 0.005 | -0.03  (-0.05, -0.01) | 0.007 |
